# Prediction of confirmed and death cases of Covid-19 in Chile through time series techniques: A comparative study

**DOI:** 10.1101/2020.12.31.20249085

**Authors:** Claudia Barría-Sandoval, Guillermo Ferreira, Katherine Benz-Parra, Pablo López-Flores

## Abstract

**Background:** Chile has become one of the countries most affected by Covid-19, a pandemic that has generated a large number of cases worldwide, which if not detected and treated in time can cause multi-organic failure and even death. The social determinants of health such as education, work, social security, housing, environment, support networks and social cohesion are important aspects to consider for the control and intervention of this pathology. Therefore, it is essential to have information about the progress of the infections at the national level and thus apply effective public health interventions. In this paper, we compare different time series methodologies to predict the number of confirmed cases and deaths from Covid-19 in Chile and thus support the decisions of health agencies;

**Methods:** We modeled the confirmed cases and deaths from Covid-19 in Chile by using ARIMA models, exponential smoothing techniques, Poisson models for time-dependent counting data. In addition, we evaluated the accuracy of the predictions by using a training set and test set;

**Results:** The database used in this paper allows us to say that for the confirmed Covid-19 cases the best model corresponds to a well-known Autoregressive Integrated Moving Average (ARIMA) time-series model, whereas for deaths from Covid-19 in Chile the best model resulted in damped trend method;

**Conclusion:** ARIMA models are an alternative to model the behavior of the spread of Covid19, however, and depending on the characteristics of the data set, other methodologies can better capture the behavior of these records, for example, Holt-winter’s method and time-dependent counting models.

## 1 Introduction

SARS-CoV-2 or also called Covid-19 is an infectious-contagious disease that has originated a pandemic of high impact on the health of the international population, causing death if not detected and treated in time.

Concerning its origin, Cruz et al. (2020) pointed out that it is still being investigated and that the virus was initially declared of zoonotic origin due to its similarity to bat coronaviruses. However, other studies Zhou et al. (2020) and De Wit et al. (2016) reveal the probable mutation of SARSCoV (transmitted to humans through consumption of exotic animals) and MERS-CoV (transmitted from camels to humans) as the etiological source of Covid-19. The most frequent symptoms cases of confirmed Covid-19 according to the World Health Organization (WHO, 2020) (https://www.who.int/es/emergencies/diseases/novel-coronavirus-2019) are fever, cough, fatigue and respiratory distress; although cases have also been reported with symtomatology atypical digestive, neurological, cardiovascular diseases, muscular diseases, ophthalmological, etc. and asymptomatic cases as indicated by Chen et al. (2020), Chang et al. (2020), Huang et al. (2020), among others.

There is no specific antiviral for the treatment of Covid-19, and several expert groups have developed guidelines for the diagnosis and treatment of SARS-CoV-2 (Jin et al., 2020). In situations where cases evolve unfavourably, some studies have pointed to the use of algorithms to address severe clinical cases (Liao et al. (2020) and Bassetti et al. (2020)). Therefore, stopping and limiting the number of infections and deaths from Covid-19 is a priority for public policy. Valdés (1995), Fernández et al. (2015) and Rebolledo et al. (2014) mentioned that poorly designed and coordinated policies can generate great biopsychosocial suffering. Therefore, health interventions must be correlated with economic and social policies to mitigate the negative impact of the pandemic in these contexts.

### 1.1 Chile and the Covid-19

The government of Chile has designed different strategies to prevent the spread of Covid-19. Among the most important measures for complying with the chilean’s health policy objectives are the border closures, cancellation of air flights, suspension of classes, suspension of non-essential supplies, social distancing, strict hygiene and personal protection measures, mandatory use of masks when interacting with other people, request for legal permits to transit or move in quarantined cities (confinement in-home or health residence), avoid touching or approaching people with respiratory infections, etc.

In the field of Chilean health, the Ministry of Health (MINSAL); establishes within its objectives to monitor the traceability of confirmed cases of Covid-19 and the real supervision of quarantines, through human resources technology. To this end, it works together with the Regional Health Services (SEREMI), which has been given the authority to supervise the traceability and isolation of cases confirmed with Covid-19. Specifically, this activity is carried out by the primary care health teams, which are made up of:

- A physician, who evaluates the clinical status of the person likely to be infected, orders the PCR test and performs an educational role for the person and his or her close contacts.
- Nursing and kinesiology professional; who has shared functions in carrying out the sample taking procedure and following up the person with probable infection using telemedicine until the PCR result is known.

If the result of the test is negative, the person is discharged; otherwise, quarantine is indicated for 14 days. After this period, the infected person is evaluated again and it is determined if he or she needs to continue in quarantine due to the persistence of the symptoms or if he or she is discharged.

The Covid-19 has exposed the social inequalities that exist in Latin America. Specifically, in Chile, according to the National Socio-Economic Characterization Survey (Encuesta CASEN, 2017, http://observatorio.ministeriodesarrollosocial.gob.cl/casen-multidimensional/casen/casen_2017.php), it is known that there is a national average of 7.3% of occupied private homes with over-crowding (5 or more people per room), which leads to greater difficulty in managing public policies. Another important social factor is the incidence of poverty (measured by family income). In Chile, according to the CASEN, 2017, this index reaches 7.4% in urban areas and 16.5% in rural areas. The Metropolitan Region has the highest percentage of households in poverty, with 26.7%, which correlates precisely with the highest number of confirmed cases of COVID 19 reported daily in the country. It is important to have a multivariate report on poverty and in this regard the Social Development Report of Social Development and Family MINDES/MDSF (MINDES/MDSF) has shown that this variable is multidimensional where it was detected in the Chilean population, through certain indicators that there are deficiencies of 22.5% in education, health, work and social security, housing and environment, as well as limitations of 10% in the networks and social cohesion, which impacts negatively and favours the increase in poverty.

### 1.2 Review of statistical models in Covid-19

Since the beginning of the pandemic, there has been a great interest in researchers and research centres for trend analysis and prediction of the rate of infections and deaths in different cities of the world. This context generates great challenges and that is why our study aims to study time series models that can describe the evolution of infections and deaths by Covid-19. The literature points out different techniques to provide accurate inferences and to establish asymptotic properties in trend estimators and time dependence of the data. One of the most popular tools for analyzing and sequentially predicting data are time series models that allow us to capture trends, breaks in structure, cycles and dependence over time. For illustrative purposes, just a few representative contributions are cited below.

In Nigeria, models of the Box-Jenkins type have been used to model and predict the propagation of Covid-19, proposed by Ibrahim and Oladipo (2020). In particular, the authors propose a model of the type Auto-Regressive Integrated Moving Average (ARIMA) of order (1,1,0) (ARIMA(1,1,0)) which provided a forecasts for ten consecutive days of the virus. Maleki et al. (2020) considered Autoregressive time series models based on two-piece scale mixture normal distributions, called TPSMNAR models for the modelling and prediction of confirmed and recovered cases of Covid-19 in the world. In the same context, Benvenuto et al. (2020) developed an ARIMA model to predict the epidemiological trend of the prevalence and incidence of Covid-19 on the Johns Hopkins epidemiological data. Liu et al. (2020) proposed a machine learning and mathematical model-based analysis by using SEIR (Susceptible, Exposed, Infectious, Recovered) and neural network models (NNs) were built to model disease trends in Wuhan, Beijing, Shanghai and Guangzhou. Combined with public transportation data, Autoregressive Integrated Moving Average (ARIMA) model was used to estimate the accumulated demands for nonlocal hospitalization during the epidemic period in Beijing, Shanghai and Guangzhou. Other models are included in the references of the following authors Perone (2020), Sarkar (2020), Tran et al. (2020) and references cited therein.

Papastefanopoulos et al. (2020) conducted a comparative study of 6-time series methods to estimate the percentage of active cases concerning the total population for ten countries with the highest number of confirmed cases from May 4, 2020. Among the methodologies proposed by these authors are the ARIMA model, HoltWinters Additive Model (HWAAS), Exponential Smoothing with Additive Trend and Additive Seasonality, Trigonometric seasonal formulation Box–Cox transformation ARMA errors and trend component (TBAT), Prophet (Automatic Forecasting Procedure), DeepAR (Probabilistic Forecasting with Auto-Regressive Recurrent Networks) and N-Beats (Neural Basis Expansion Analysis for Interpretable Time Series Forecasting). These authors use the root mean square error (RMSE) to assess the performance of each time series model, concludes that traditional statistical methods such as ARIMA and TBAT, in general, prevail over their deep learning counterparts such as DeepAR and N-BEATS and argue that the result is not a surprise given the lack of large amounts of data.

In the same line, Yonar et al. (2020) proposed a study to predict and model the number of Covid-19 cases using two methodologies: ARIMA and exponential smoothing methods. In this paper, the authors mention that for different countries under study there is no single model to describe the behaviour of the number of cases, but according to the characteristics of the data, both methods are effective in describing the virus propagation curves. Other methods have been studied, for example, in Agosto and Giudici (2020) a regression Poisson autoregressive model is proposed to understand Covid-19 contagion dynamics, Mizumoto and Chowell (2020) fitted the reported serial interval (mean and standard deviation SD) with a gamma distribution and applied the `“earlyR”` package in `R` (Team (2017)) to estimate *R*0 in the early stage of the Covid-19 outbreak. Finally, Roda et al. (2020) written a paper that emphasizes the difficulty of modeling the characteristics of the Covid19 cases. These authors used SIR and SEIR models for model predictions and applied model-selection analysis. Following the investigations of authors above mentioned, we will analyze other methodologies to contribute to the discussion of the current topic. In particular, in this paper, a comparative analysis between the ARIMA models, exponential smoothing, space state models, Bayesian approach and GLARMA model are proposed. The last three methods are used through a Poisson distribution for counting data with local linear trend model.

The article is organized as follows. In Section 2, a brief description of the data set and the time series models used in this paper are described in detail. In Section 3, the behavior of the differents models is examined by means of a statistical analysis which includes estimations of the parameters and goodness of fit of the residuals of each model. The main conclusions are summarized in Section 4.

## 2 Materials and Methods

### 2.1 Dataset

In this paper we will carry out an analysis of the number of confirmed cases and deaths by Covid-19 in Chile from March 2, 2020 to July 14, 2020. The data was obtained from the Ministry of Science and Technology, Knowledge and Innovation http://www.minciencia.gob.cl/covid19. Figure 1(a) displays the number of confirmed cases of Covid-19 in Chile, where a peak of infections is observed on June 15, which drops to July 14. Furthermore, Figure1 (b) shows the deaths from Covid-19, where an exponential growth is observed up to the date of this study.

**Figure 1:**
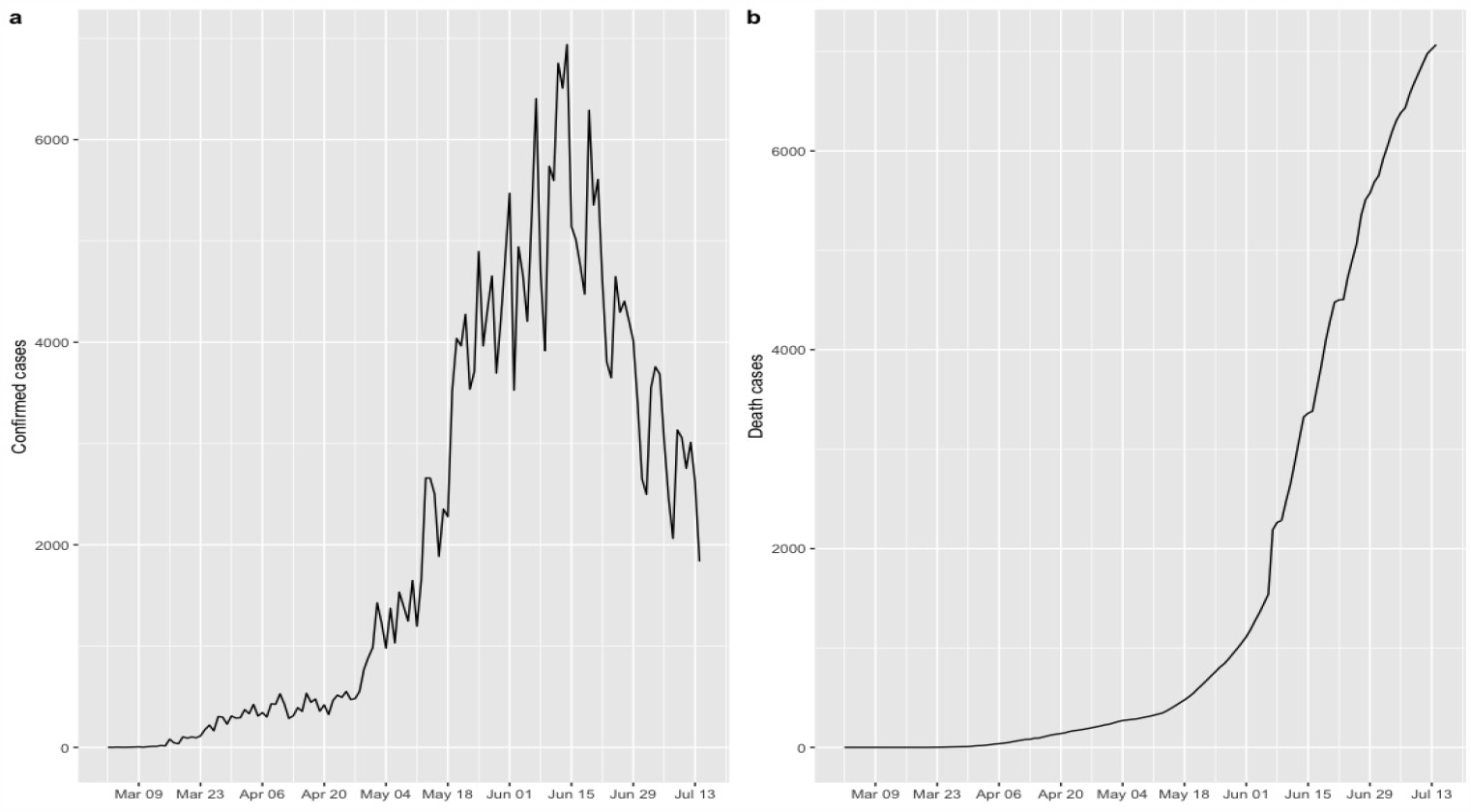
Covid-19 in Chile. (a) Confirmed cases. (b) Deaths cases

### 2.2 Model development

Time series models are a very effective tool for modelling data recorded sequentially over time. The objective of this methodology is to capture the time dependence between observations through a mathematical model that allows the description of the main characteristics of the data. In general, these records present trends and seasonal components which can be modelled by different statistical techniques. In what follows, a description of the most used models in time series will be made; such as ARIMA(*p, d, q*) processes, a random walk with trend for count data, among others.

- ARIMA Model

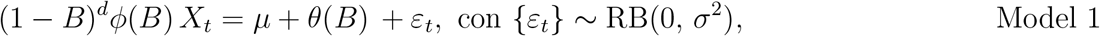

where *d* is the positive integer parameter of integration, *ϕ*(*z*) = 1 − *ϕ*_1_ *z*−… − *ϕ*_*p*_ *z*^*p*^ autoregressive polynomial and *θ*(*z*) = 1+*θ*_1_ *z*+… +*θ*_*p*_ *z*^*q*^ moving-average polynomial. Ibrahim and Oladipo (2020) used this model to predict the propagation of Covid-19 in Nigeria, for more details of this model the reader is referred to the time series book written by Brockwell and Davis (1991).
- Poisson Model Another model for working with time-series count data is the Poisson process with a local linear trend model defined by

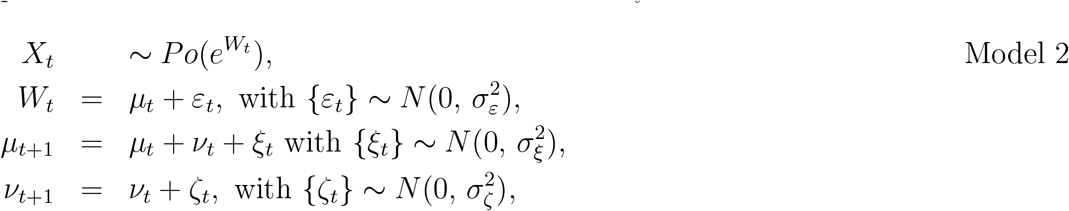

where *µ*_*t*_ is a random walk with a drift component given by *ν*_*t*_ and {*ε*_*t*_} is a white noise process which captures the extra variations of the time series. The Model 2 estimates and predictions will be made through two methodologies that are widely discussed in the literature (Durbin and Koopman (2012), Robert and Casella (2013) among other authors), namely
  a. Space-state models
  b. Bayesian analysis.
  For Model 2b) we consider a random walk of order 1, i.e.,

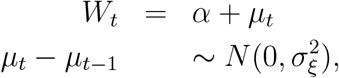

where 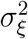 is estimated as precision 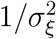 in Bayesian framework.
- GLARMA(*p, q*) Model On the other hand, Dunsmuir (2015) provided other methology for account data with serial dependence in regression modeling of time series called generalized linear autoregressive moving average (GLARMA(*p, q*)) models defined by

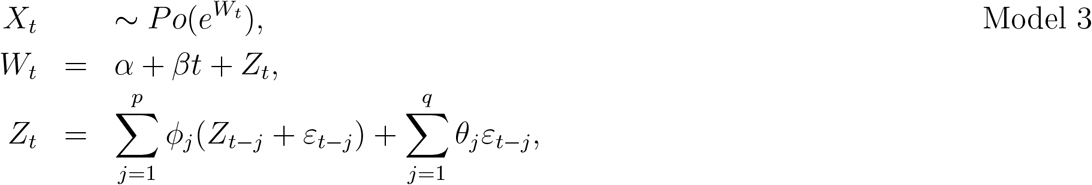

where *ε*_*t*_ are defined as 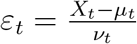, with 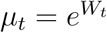 and *ν*_*t*_ is some scaling sequence.
- Holt’s local trend and Damped trend method Finally, Holt (1957) and Winters (1960) proposed a methodology known as Holt-Winters method. This method is a more general class than the exponential smoothing method which explains the level and trend as follows

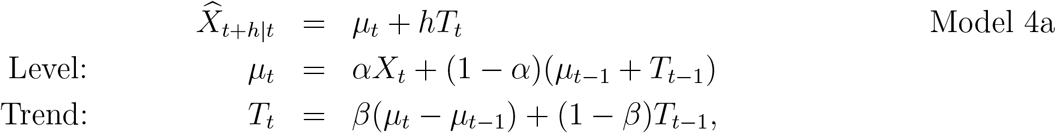

where 0 ≤ *α* ≤ 1 is the smoothing parameter, 0 ≤ *β* ≤ 1 is the smoothing parameter for the trend. The *h*-step ahead forecasts 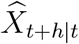 are calculated using the smoothing equations for the level *µ*_*t*_ and the trend *T*_*t*_. Gardner Jr and McKenzie (1985) have developed an exponential smoothing model designed to damp erratic trends defined as follows

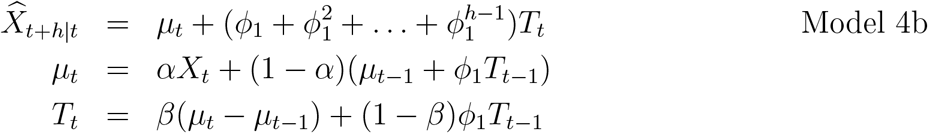

where 0 < *ϕ*_1_ < 1 is damping parameter. Its dampens the trend to be more conservative for longer forecast horizons.

## 3 Result and Discussion

In this section, we will analyze the performance of Models 1-4 described in Section 2.2. Again the data consists of the number of confirmed and deaths by Covid-19 in Chile from March 2, 2020 to July 14. For the estimations, we used the `R` free software (Team (2017)). Table 1 shows the parameter estimates and the estimated SD (in parentheses) of the Model 1-4 for both time series (confirmed and deaths cases). For Model 1, an ARIMA(0, 1, 2) model with a drift to confirmed cases and an ARIMA(0, 2, 3) model with-out drift to death cases is proposed. `Arima` command from the `forecast` package, whereas `KFAS` package (Helske (2016)) is used for Model 2a) and integrated nested Laplace approximation (INLA) provided by the package `INLA` (Lindgren et al. (2015)) for the estimation of Model 2b). The estimates of Model 3 are based in `GLARMA` package (Dunsmuir et al., 2015). For the confirmed cases data we propose a GLARMA(2, 0) whereas for death data a GLARMA(1, 0) is propose, both models with *ν*_*t*_ = 1. Additionally, we use the `holt` command from the `forecast` package to estimate the parameters of Model 4a and 4b by adding the argument `damped=TRUE`. Figures 2-3 displays the fitted values for each Model, the dashed lines represents the actual data, while continuous line represents the fitted values which are shown in coloured curves for Models 1-4. From these figures, we can conclude that the best model that captures the trend and time dependence characteristics are Models 2. Figure 4-5 presents the sample autocorrelation function (ACF) and partial autocorrelation function (PACF) of the residuals of the estimated models for both data set. From these Figures, it seems that there in no significant autocorrelations in the residuals, except for the Model 3. In what follows, we will evaluate the predictive power of the proposed models.

**Table 1:**
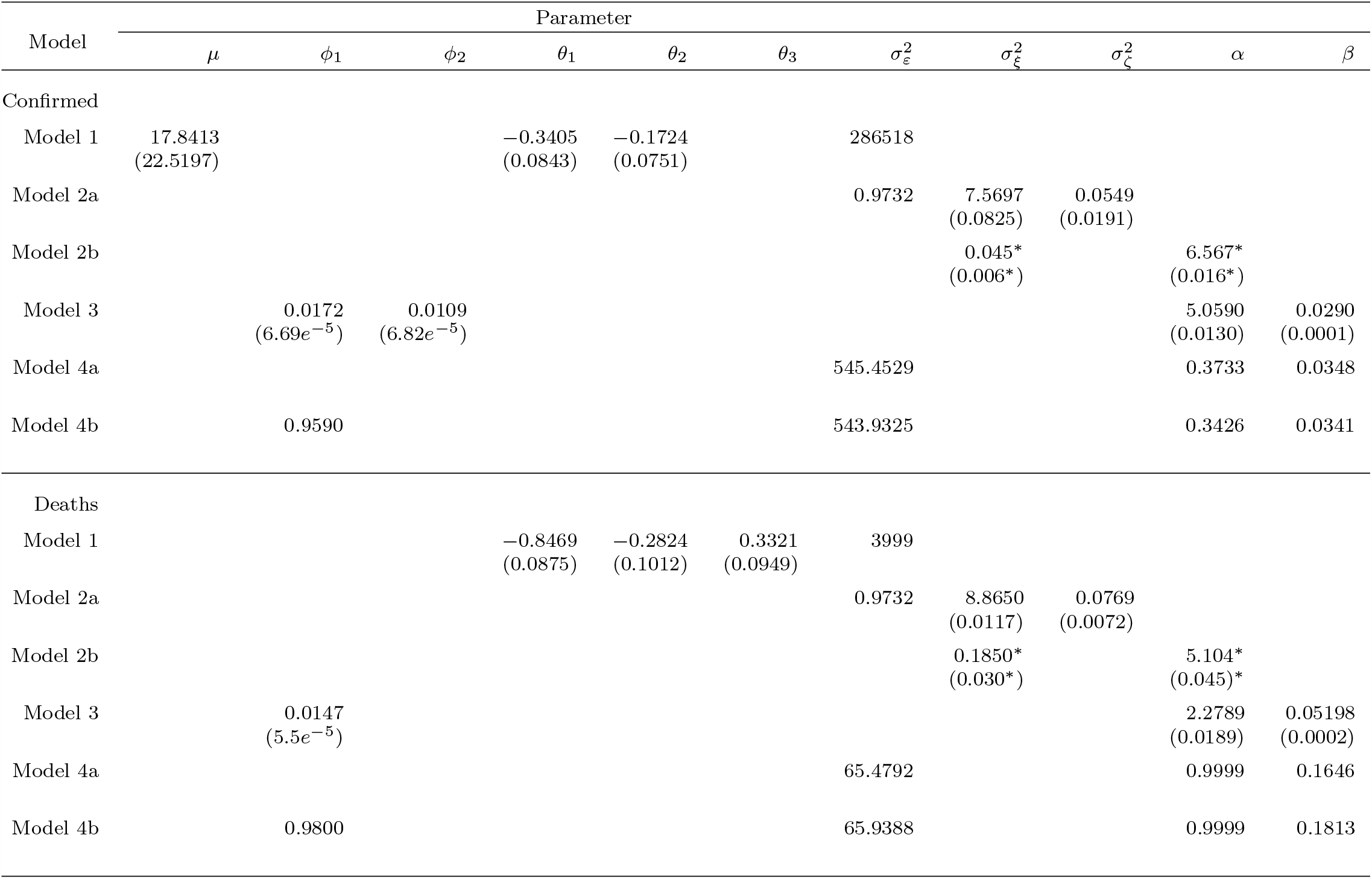
Table reports the estimated parameters of the Models 1-4 on the Covid-19 series. The table also reports posterior mean and SD of the parameters from fitting Model 2b.

**Figure 2:**
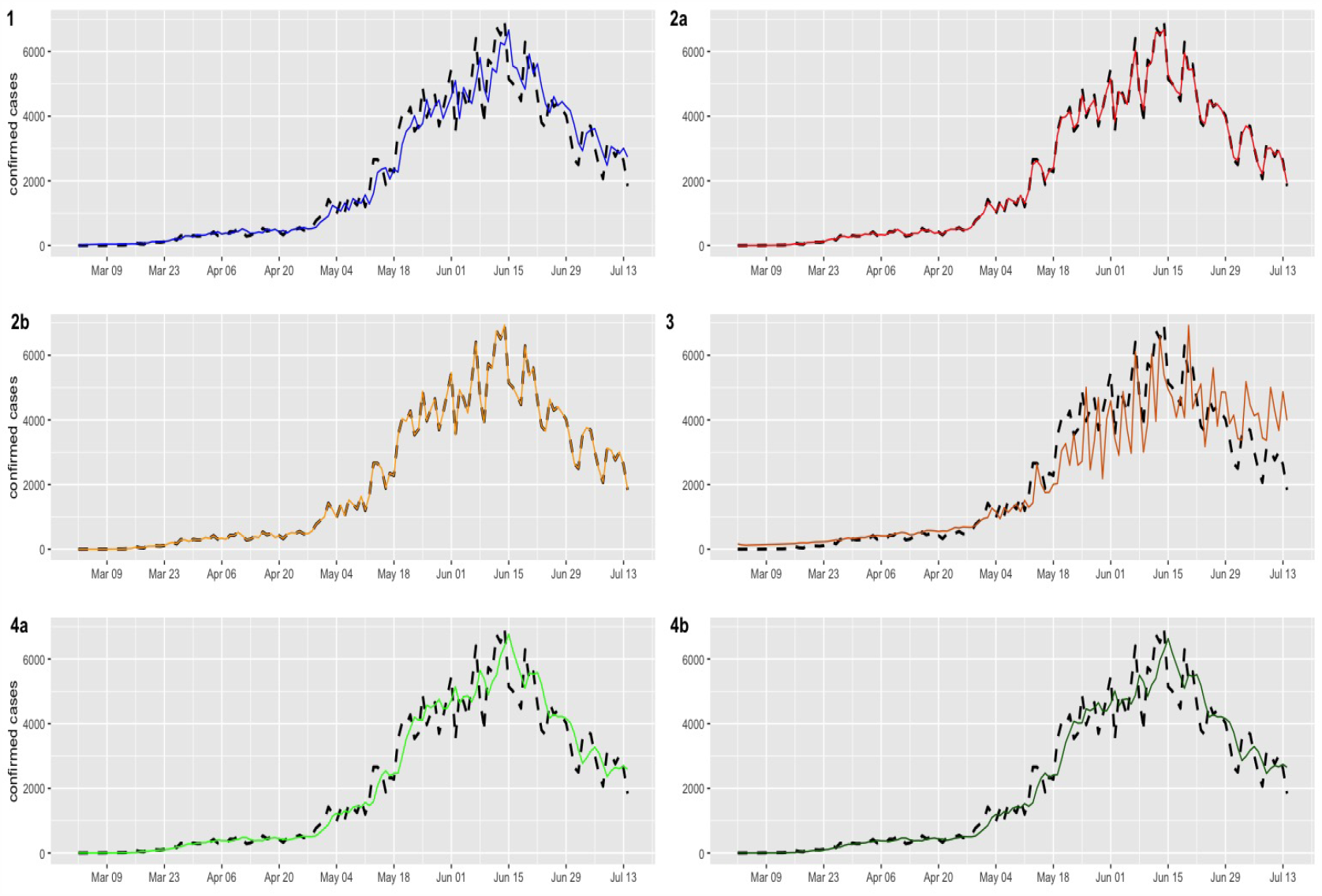
Confirmed cases (black dashed lines) versus fitted values (continuous line)

**Figure 3:**
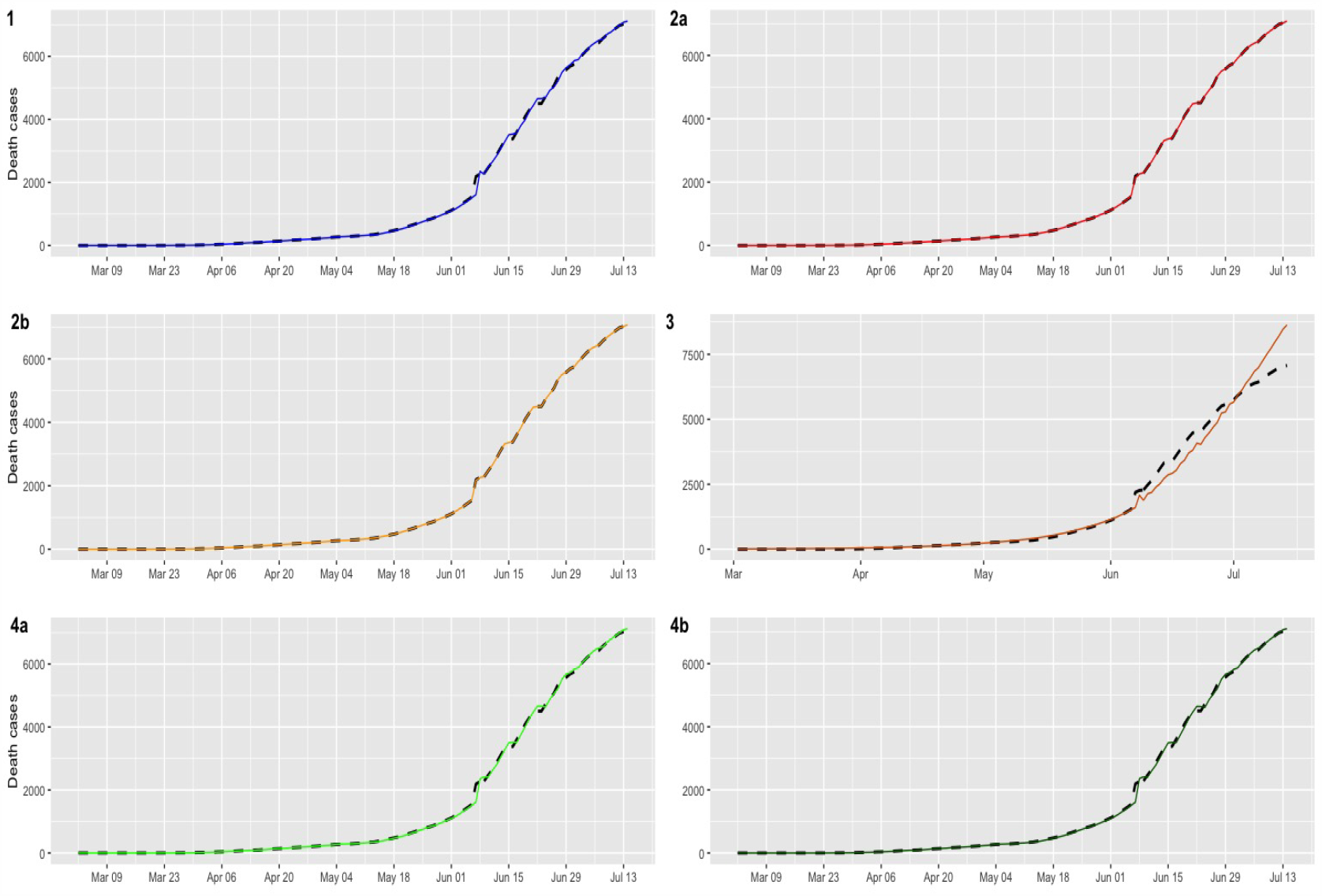
Deaths cases (black dashed lines) versus fitted values (continuous line)

**Figure 4:**
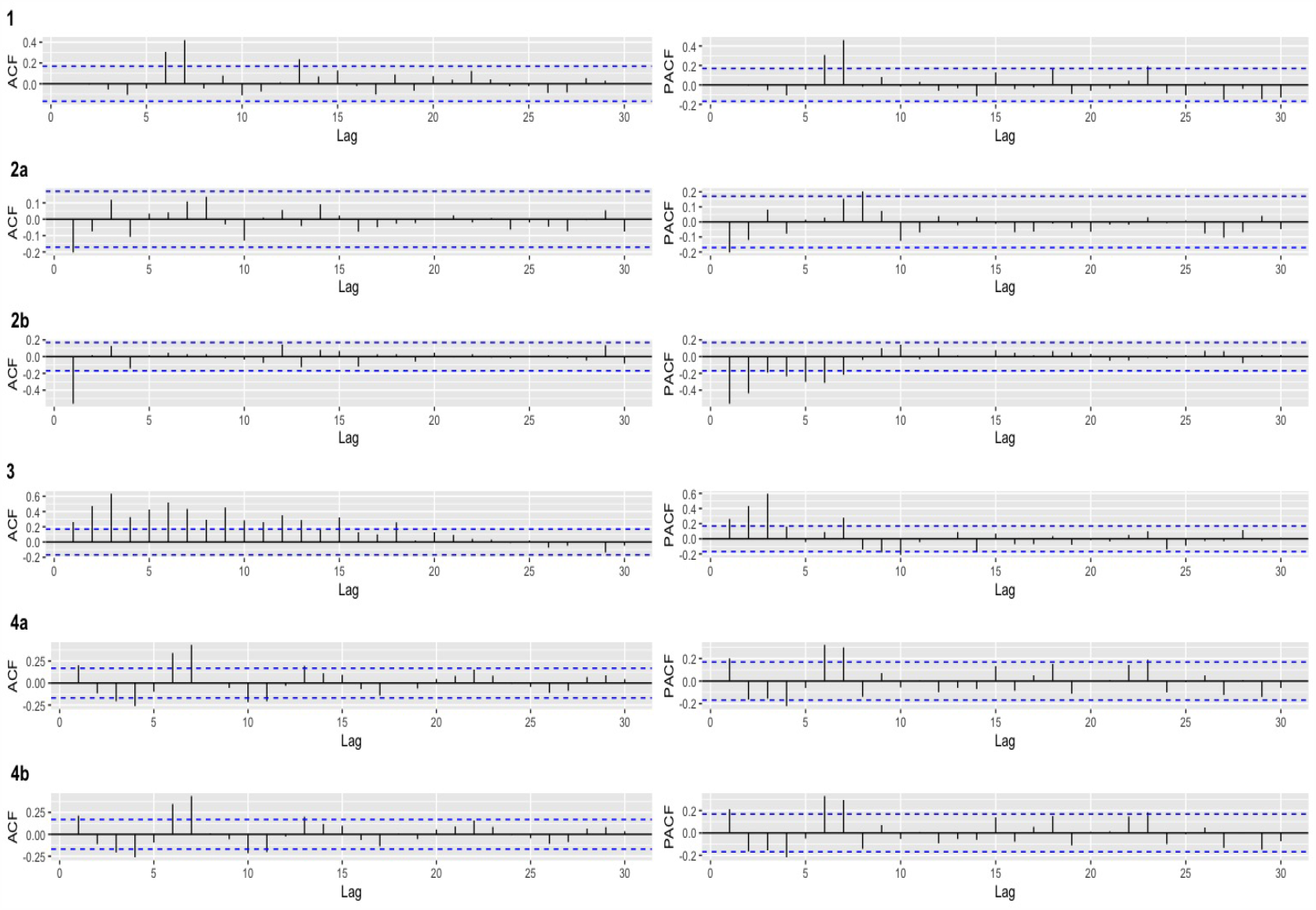
Confirmed Covid-19 cases: ACF and PACF plots of the residuals for the fitted models.

**Figure 5:**
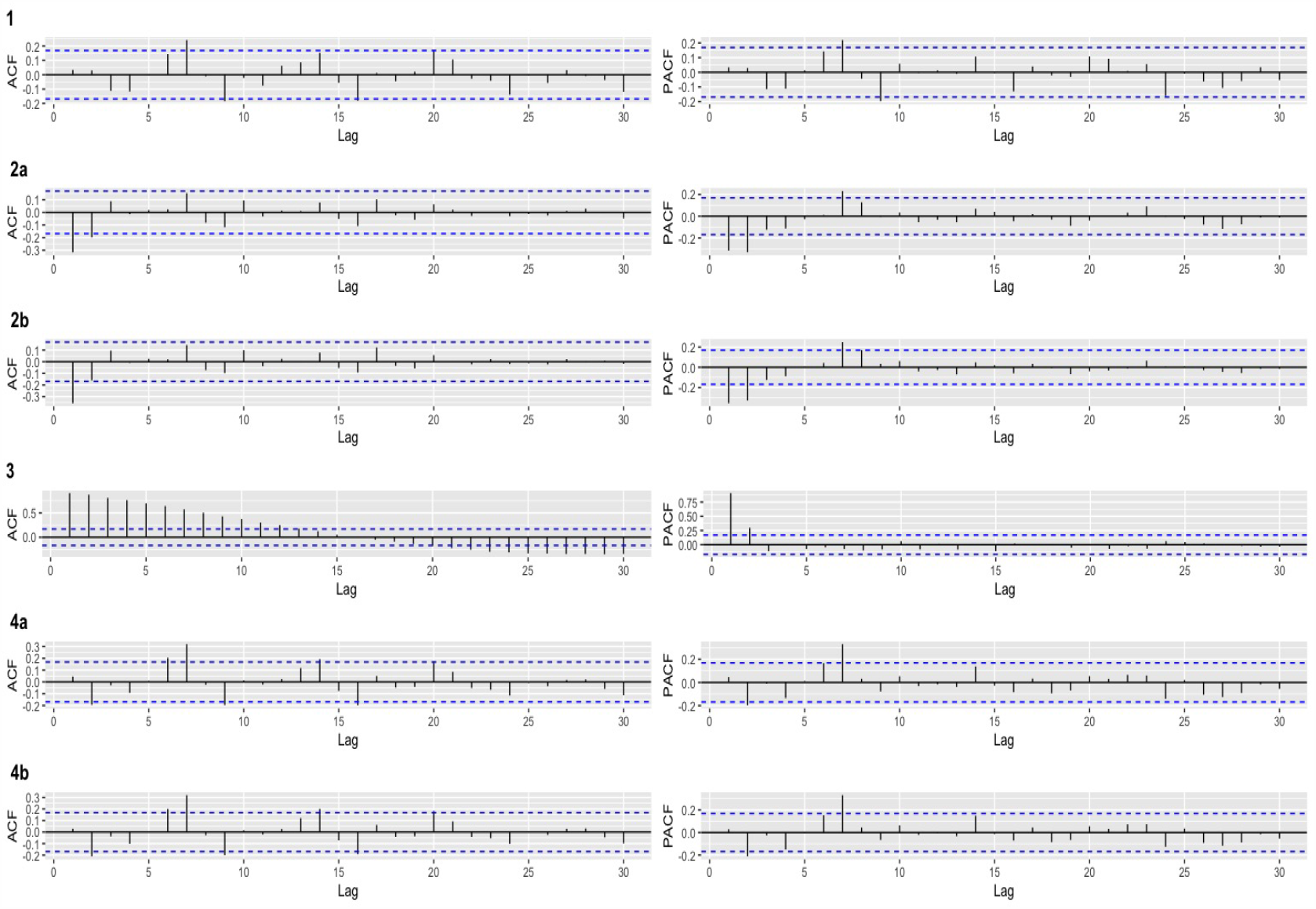
Deaths Covid-19 cases: ACF and PACF plots of the residuals for the fitted models.

### 3.1 Analysis of ex post forecast accuracy

In this section, we evaluate the accuracy of the predictions by using a training set and test set. We consider a training set {*X*_*t*_} from March 02 to July 06 (estimation period) with a total of *N* = 129 observations and test data {*X*_*t*_} from July 07 to July 14 (validation period) which will be used to *m*-step-ahead prediction with *m* = 6 daily observations. The values 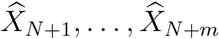 are called ex post forecast or period forecast with the starting period July 07. The *m*-step-ahead forecasts are compared with the validation period giving rise ex post forecast errors, i.e. 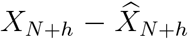 for horizont *h* = 1, …, *m*. The errors was assessed by the statistics of residuals such as; Mean error (ME), Root mean squared error (RMSE), Mean absolute error (MAE), Mean percentage error (MPE) and Mean absolute percentage error (MAPE). Table 2 reports the statistics of ex post forecast errors of the models in both dataset. In the case of confirmed cases all indicators in favor of the ARIMA model, i.e., Model 1 with a MAPE of the 17.5% and for the deaths Covid-19 cases the statistics favor to damped trend method, i.e., Model 4b with MAPE equal to 0.37% which is reasonably low values demonstrating the suitability of the proposed model for prediction. The above is supported by the Figures 6-7.

**Table 2:** The descriptive statistics of ex post forecast errors

| Model | ME | RMESE | MAE | MPE | MAPE |
| --- | --- | --- | --- | --- | --- |
| Confirmed cases |  |  |  |  |  |
| Model 1 | -222.3229 | 510.1141 | 454.268 | 4.903861 | 17.53703 |
| Model 2a | -164.3078 | 838.1199 | 614.4429 | -12.22922 | 26.74773 |
| Model 2b | 422.8959 | 693.6704 | 643.8252 | 11.81886 | 23.85205 |
| Model 3 | -1764.664 | 1904.865 | 1764.664 | -69.39226 | 69.39226 |
| Model 4a | 735.086 | 784.1449 | 735.086 | 25.87431 | 25.87431 |
| Model 4b | 551.5119 | 635.8757 | 587.9894 | 18.47044 | 20.45723 |
| Deaths cases |  |  |  |  |  |
| Model 1 | -56.8675 | 75.45366 | 56.8675 | -0.81314 | 0.81314 |
| Model 2a | -2018.527 | 2320.163 | 2018.527 | -28.93521 | 28.93521 |
| Model 2b | 138.3735 | 145.9142 | 138.3735 | 1.992866 | 1.992866 |
| Model 3 | -2891.794 | 2900.363 | 2891.794 | -41.84812 | 41.84812 |
| Model 4a | -83.45833 | 109.3747 | 83.45833 | -1.191752 | 1.191752 |
| Model 4b | -16.01247 | 39.42867 | 26.08101 | -0.22479 | 0.371870 |

**Figure 6:**
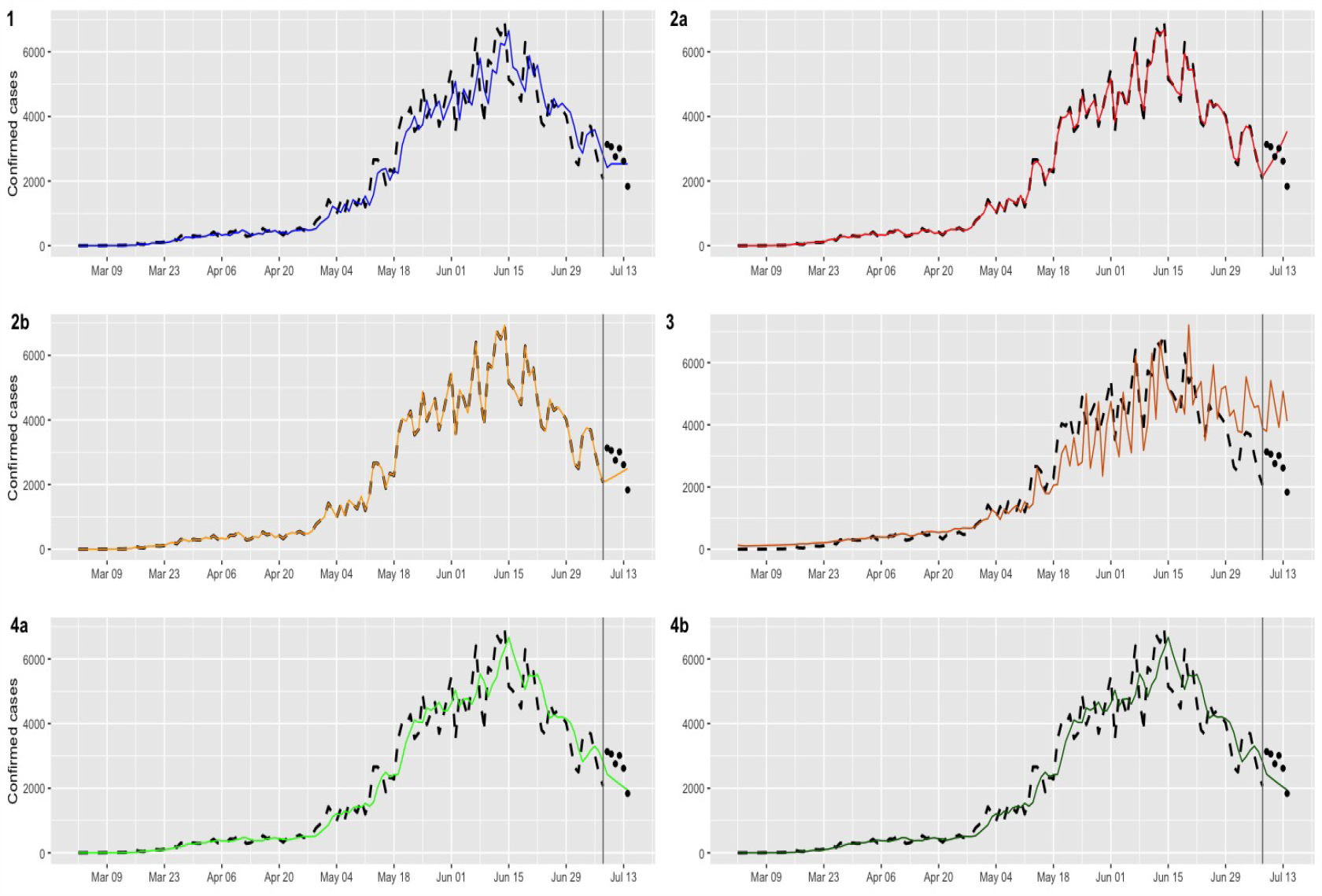
Multi-step forecasts for confirmed Covid-19 cases (black dashed lines). Continuous line and dots represents fitted values and ex post forecast respectively

**Figure 7:**
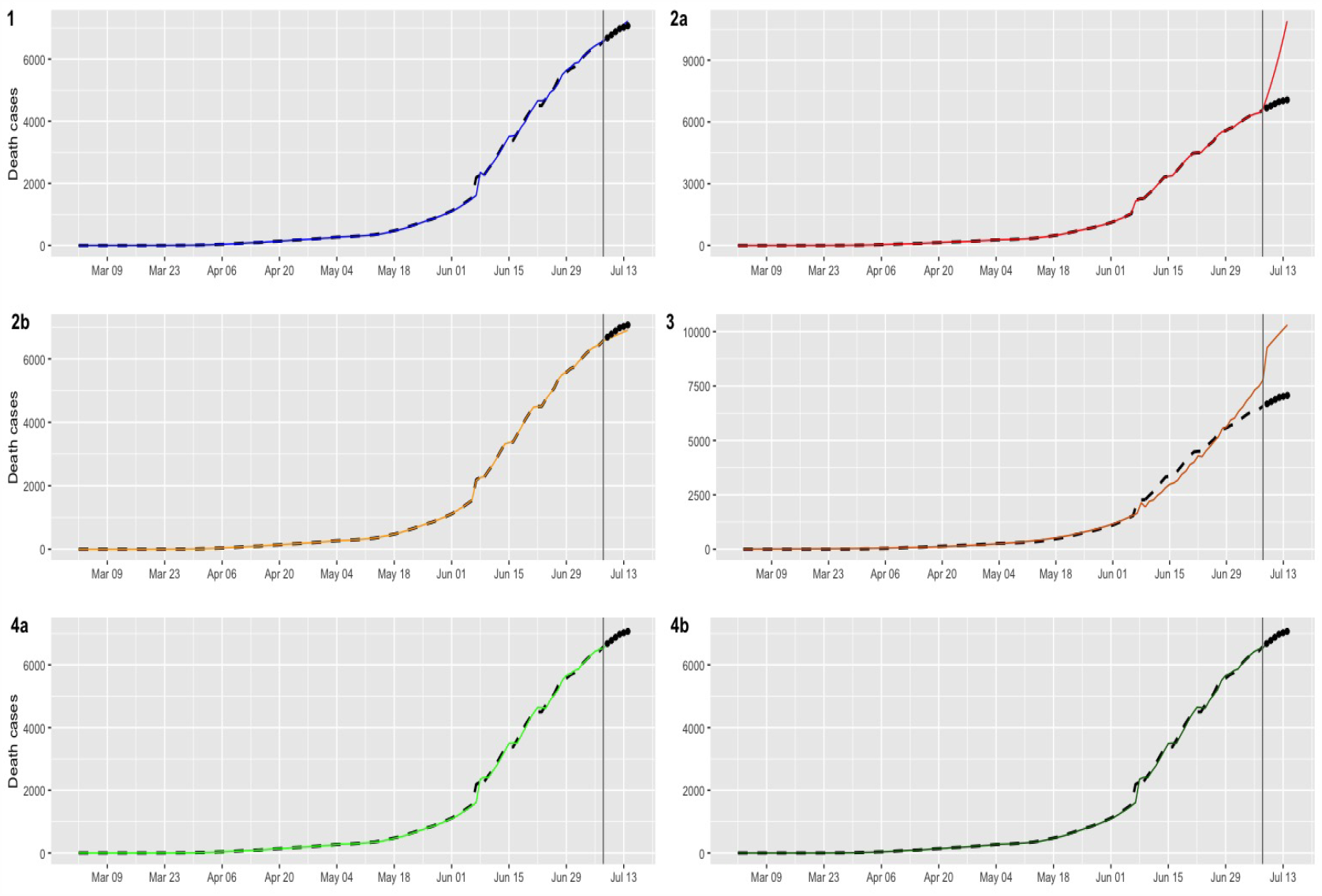
Multi-step forecasts for deaths Covid-19 cases (black dashed lines). Continuous line and dots represents fitted values and ex post forecast respectively

## 4 Conclusions

In this paper we have proposed a comparative analysis of the most used time series models in sequential data modeling. In particular, an ARIMA model, space-state model, a Bayessian model for counting data, and exponential smoothing techniques were proposed. The main motivation of this paper is to contribute to the discussion on the mathematics models to provide predictions of confirmeds and deaths cases related to the Covid-19 and thus have relevant information for a timely decision taken by the Government of Chile.

The database used in this paper allows us to say that for the confirmed Covid-19 cases the best model corresponds to a well-known Autoregressive Integrated Moving Average (ARIMA) time-series model, whereas for deaths from Covid-19 in Chile the best model resulted in damped trend method. In line with other authors, we can affirm that the proposal of this paper does not imply its global use in the prediction of confirmed cases and deaths from Covid-19, since the performance of this model is subject to the biopsychosocial determinants of each country. It would be interesting, in a future work, to develop Machine Learning techniques to model the behavior of these curves, subject to the availability of large volumes of data and carry out a statistical study to correlational the spread of the virus to the Chileans biopsychosocial determinants.

## Data Availability

The data was obtained from the Ministry of Science and Technology, Knowledge and Innovation of Chile.

http://www.minciencia.gob.cl/covid19

## References

1. Agosto, A. and P. Giudici (2020). A poisson autoregressive model to understand covid-19 contagion dynamics. Available at SSRN 3551626.

2. Bassetti, M., A. Vena, and D. R. Giacobbe (2020). The novel chinese coronavirus (2019-ncov) infections: Challenges for fighting the storm. European journal of clinical investigation 50 (3), e13209.

3. Benvenuto, D., M. Giovanetti, L. Vassallo, S. Angeletti, and M. Ciccozzi (2020). Application of the arima model on the covid-2019 epidemic dataset. Data in brief, 105340.

4. Brockwell, P. J. and R. A. Davis (1991). Time Series: Theory and Methods (Second ed.). New York: Springer.

5. Chang, D., M. Lin, L. Wei, L. Xie, G. Zhu, C. S. D. Cruz, and L. Sharma (2020). Epidemiologic and clinical characteristics of novel coronavirus infections involving 13 patients outside wuhan, china. Jama 323 (11), 1092–1093.

6. Chen, L., H. Liu, W. Liu, J. Liu, K. Liu, J. Shang, Y. Deng, and S. Wei (2020). Analysis of clinical features of 29 patients with 2019 novel coronavirus pneumonia. Zhonghua jie he he hu xi za zhi= Zhonghua jiehe he huxi zazhi= Chinese journal of tuberculosis and respiratory diseases 43, E005–E005.

7. Cruz, M. P., E. Santos, M. V. Cervantes, and M. L. Juaárez (2020). Covid-19, a worldwide public health emergency. Revista Clínica Espanñola (English Edition).

8. De Wit, E., N. Van Doremalen, D. Falzarano, and V. J. Munster (2016). Sars and mers: recent insights into emerging coronaviruses. Nature Reviews Microbiology 14 (8), 523.

9. Dunsmuir, W. T. (2015). Generalized linear autoregressive moving average models. Handbook of Discrete-Valued Time Series. CRC Monographs.

10. Dunsmuir, W. T., D. J. Scott, et al. (2015). The glarma package for observation-driven time series regression of counts. Journal of Statistical Software 67 (7), 1–36.

11. Durbin, J. and S. J. Koopman (2012). Time series analysis by state space methods. Oxford university press.

12. Fernández, P. G., F. C. Solar, S. S. Bórquez, and C. B. Navarrete (2015). Psychometric analysis and adaptation of the social distance scale (ds) in a chilean sample. Salud Mental 38 (2), 117–122.

13. Gardner Jr, E. S. and E. McKenzie (1985). Forecasting trends in time series. Management Science 31 (10), 1237–1246.

14. Helske, J. (2016). Kfas: Exponential family state space models in r. arXiv preprint arXiv:1612.01907.

15. Holt, C. C. (1957). Forecasting trends and seasonal by exponentially weighted moving averages. ONR Memorandum 52.

16. Huang, C., Y. Wang, X. Li, L. Ren, J. Zhao, Y. Hu, L. Zhang, G. Fan, J. Xu, X. Gu, et al. (2020). Clinical features of patients infected with 2019 novel coronavirus in wuhan, china. The lancet 395 (10223), 497–506.

17. Ibrahim, R. R. and O. H. Oladipo (2020). Forecasting the spread of covid-19 in nigeria using box-jenkins modeling procedure. medRxiv.

18. Jin, Y.-H., L. Cai, Z.-S. Cheng, H. Cheng, T. Deng, Y.-P. Fan, C. Fang, D. Huang, L.-Q. Huang, Q. Huang, et al. (2020). A rapid advice guideline for the diagnosis and treatment of 2019 novel coronavirus (2019-ncov) infected pneumonia (standard version). Military Medical Research 7 (1), 4.

19. Liao, X., B. Wang, and Y. Kang (2020). Novel coronavirus infection during the 2019–2020 epidemic: preparing intensive care units?the experience in sichuan province, china. Intensive care medicine 46 (2), 357–360.

20. Lindgren, F., H. Rue, et al. (2015). Bayesian spatial modelling with r-inla. Journal of Statistical Software 63 (19), 1–25.

21. Liu, Z., S. Huang, W. Lu, Z. Su, X. Yin, H. Liang, and H. Zhang (2020). Modeling the trend of coronavirus disease 2019 and restoration of operational capability of metropolitan medical service in china: a machine learning and mathematical model-based analysis. Global Health Research and Policy 5, 1–11.

22. Maleki, M., M. R. Mahmoudi, D. Wraith, and K.-H. Pho (2020). Time series modelling to forecast the confirmed and recovered cases of covid-19. Travel Medicine and Infectious Disease, 101742. MINDES/MDSF. Informe de desarrollo social, 2019.

23. Mizumoto, K. and G. Chowell (2020). Transmission potential of the novel coronavirus (covid-19) onboard the diamond princess cruises ship, 2020. Infectious Disease Modelling.

24. Papastefanopoulos, V., P. Linardatos, and S. Kotsiantis (2020). Covid-19: A comparison of time series methods to forecast percentage of active cases per population. Applied Sciences 10 (11), 3880.

25. Perone, G. (2020). An arima model to forecast the spread and the final size of covid-2019 epidemic in italy. medRxiv.

26. Rebolledo, E. L. R. Mesí a, and G. Silva (2014). Nonattendance to medical specialists appointments and its relation to regional environmental and socioeconomic indicators in the chilean public health system. Medwave 14 (9), e6023–e6023.

27. Robert, C. and G. Casella (2013). Monte Carlo statistical methods. Springer Science & Business Media.

28. Roda, W. C., M. B. Varughese, D. Han, and M. Y. Li (2020). Why is it difficult to accurately predict the covid-19 epidemic? Infectious Disease Modelling.

29. Sarkar, D. (2020). Covid 19 pandemic: A real-time forecasts & prediction of confirmed cases, active cases using the arima model & public health in west bengal, india. medRxiv.

30. Team, R. C. (2017). R core team (2017). r: A language and environment for statistical computing. R Found. Stat. Comput. Vienna, Austria.

31. Tran, T., L. Pham, and Q. Ngo (2020). Forecasting epidemic spread of sars-cov-2 using arima model (case study: Iran). Global Journal of Environmental Science and Management 6 (Special Issue (Covid-19)), 1–10.

32. Valdés, J. G. (1995). Pinochet’s economists: The Chicago school of Economics in Chile. Cambridge University Press.

33. WHO, C. (2020). consultado 2 feb 2020.

34. Winters, P. R. (1960). Forecasting sales by exponentially weighted moving averages. Management Science 6 (3), 324–342.

35. Yonar, H., A. Yonar, M. A. Tekindal, and M. Tekindal (2020). Modeling and forecasting for the number of cases of the covid-19 pandemic with the curve estimation models, the box-jenkins and exponential smoothing methods. EJMO 4 (2), 160–165.

36. Zhou, P., X.-L. Yang, X.-G. Wang, B. Hu, L. Zhang, W. Zhang, H.-R. Si, Y. Zhu, B. Li, C.-L. Huang, et al. (2020). A pneumonia outbreak associated with a new coronavirus of probable bat origin. nature 579 (7798), 270–273.

